# Mixed invasive molds among COVID-19 patients

**DOI:** 10.1101/2021.08.09.21261555

**Authors:** Vanya Singh, Amber Prasad, Prasan Kumar Panda, Manjunath Totaganti, Amit Tyagi, Abhinav Thaduri, Shalinee Rao, Mukesh Bairwa, Ashok Kumar Singh

## Abstract

**Purpose:** Due to surge in COVID cases during the second wave of the COVID pandemic, the healthcare system collapsed in India with shortage of hospital beds, injudicious use of steroids and other immunomodulators, and poor glycaemic monitoring among a population with pre-existing risk of diabetes. Fungal epidemic was announced amid COVID pandemic with several cases of COVID-associated mucormycosis and aspergillosis being reported. But, there is no data regarding mixed fungal infections in COVID patients.

**Materials and Methods:** The study presented a series of ten consecutive cases with dual invasive molds in patients infected with SARS-CoV-2. Among patients hospitalized with the diagnosis of COVID in May 2021 at a tertiary care center in North India, ten microbiologically confirmed dual/mixed COVID-associated mucor-aspergillosis (CAMA) were evaluated.

**Results:** All patients were diabetics with the majority having severe COVID pneumonia (6/10, 60%) either on admission or in the past one month, whilst two were each of moderate (20%) and mild (20%) categories of COVID. The patients were managed with amphotericin-B along with surgical intervention. In this case series, 70% of all CAMA (*Rhizopus arrhizus* with *Aspergillus flavus* in seven and *Aspergillus fumigatus* in three patients) patients survived, connoting the critical importance of a high index of clinical suspicion and accurate microbiological diagnosis for managing invasive molds.

**Conclusions:** Mixed fungal infections i.e. CAMA during COVID and post-COVID periods may be an emerging disease. This outbreak is seen particularly in such patients with uncontrolled diabetes, on steroids, or cocktail therapy, or living in unhygienic environments.We believe that our findings would help gain a better insight into the risk and progression of invasive fungal mixed infections among COVID patients and thus play a pivotal role in diagnosing, classifying, and implementing an effective management strategy for treating similar cases in the future.

## INTRODUCTION

Over the past one and half years, the severe acute respiratory syndrome coronavirus-2 (SARS-CoV-2) has presented with a myriad of manifestations and complications [1,2]. Whilst the world managed to recover from the first wave of COVID-19, the second wave has put the world at wits end, with the SARS-CoV-2 virus bringing up new surprises and tricks. The second wave has especially hit hard on India, especially the recent increased incidence of invasive fungal infections.

The usually rare acute invasive fungal rhinosinusitis cases have surged in post-COVID patients [3-5]. Since there is a lack of population-based studies in the literature to estimate the baseline incidence prevalence rate of mucormycosis in India, it is difficult to state that the overall incidence rate of COVID-associated mucormycosis (CAM) has increased [6]. COVID-associated pulmonary aspergillosis (CAPA) is another entity that came into the picture during the first wave of the COVID pandemic [7]. But mixed/dual invasive fungal infections associated with COVID-19 have rarely been reported.

To the best of our knowledge, we report the first case series of ten consecutive cases of COVID-associated mucor-aspergillosis (CAMA) with their associated risk factors, clinical presentation, diagnosis, treatment, and outcome.

## METHODS

### Study Design and patients

This case series is part of a project titled disease profile of COVID-l9 including their follow-up in patients at a tertiary institute in India, approved by the Institutional Ethical Committee, All India Institute of Medical Sciences (AIIMS), Rishikesh, India (CTRI/2020/08/027169). Baseline demographics, presenting signs and symptoms, disease characteristics, microbiological and radiological findings, treatments, and mortality outcomes of the study population were noted.

### Inclusion criteria

Among all patients who were either known case of previous COVID infection or whose reverse transcriptase polymerase chain reaction test (RT-PCR) for COVID came positive after admission during May 2021 and presented with one or more of the following symptoms: decreased/loss of vision, sinusitis, headache, facial cellulitis, diplopia, proptosis, toothache, loosening of teeth, blackish discoloration over the bridge of nose/palate, prolonged fever, jaw involvement, altered mental status, necrosis of tissue with black crusts; ten consecutive patients whose nasal tissue sample showed both broad aseptate ribbon-like hyphae with perpendicular branching and acute-angled thin hyaline septate hyphae on potassium hydroxide (KOH) mount or whose fungal culture reports showed pure growth of both *Mucorales* as well as *Aspergillus* species were included in the study.

### Procedures

A detailed history, comprehensive otorhinolaryngological, ophthalmologic evaluation, and neurological examinations were made to determine the disease’s extent and severity. Routine blood investigations, including complete blood counts, blood sugar and liver function tests, kidney function tests, along with HbA1C, serum procalcitonin, serum ferritin, were done.

Microscopy (direct and/or histopathology) and culture are the cornerstones of diagnosis. Scraping or exudate from the nasal cavity and/or paranasal sinuses, hard palatal lesions, sinus material, a biopsy from extracted tooth socket area, endoscopic collection of debrided tissue/biopsy, etc were sampled [6]. Diagnosis of mixed fungal infections was made if two types of fungal hyphae were visible on direct sample KOH. Demonstration of broad aseptate ribbon-like hyphae with right-angled branching on 10% KOH preparation of specimens obtained and Sabouraud’s Dextrose Agar (SDA) culture reports was suggestive of *Mucorales*. In contrast, demonstration of acute-angled thin hyaline septate hyphae was suggestive of *Aspergillus* spp. Lactophenol cotton blue (LPCB) was used to stain fungal elements obtained on culture growth [8]. Histopathological examination (HPE) was also done for the specimens taken during surgery, like sinus, maxilla, orbital fat etc. H&E and PAS stains were used to confirm presence of fungal hyphae.

Radiological investigations, including CT and/or MRI of paranasal sinuses, orbit, brain, and thorax, were obtained to identify the disease’s extent. The patients were diagnosed as proven, probable, or possible cases of invasive fungal diseases (IFD), based on European Organisation for Research and Treatment of Cancer and the Mycosis Study Group Education and Research Consortium definitions and ECMM/ISHAM consensus criteria [8,9]. The patients were given medical and/or surgical management depending on the extent of their disease, and a mutual decision for management was undertaken by a multidisciplinary team [10].The outcome for treatment was assessed in terms of the success and failure of management. Treatment success was defined as a stable and disease-free patient or stable patient on follow-up. Treatment failure was defined as the death of the patient due to fungal invasion.

## RESULTS

The main demographic, clinical characteristics, and associated risk factors for fungal infections of the ten patients at the time of hospital admission were described (Table 1). Only six were confirmed COVID positive before admission. The remaining four cases were never tested for COVID before admission despite having clinical symptoms suggestive of the same and were diagnosed with COVID by RT-PCR after admission. The major events of the course of illness for each case were presented (Figure 1). There were six men and four women with a mean age of 49.2 ± 8.8 years, with a minimum age of 34 and maximum age of 62 years. All patients had history of COVID within one month (15-23 days) of presentation (Figure 1, Table 1) and were diabetics. Eight had uncontrolled diabetes mellitus with poor glycaemic control (HbA1C >7.0), one was pre-diabetic, and one had long-duration diabetes (>7 years) with glycaemic control (Table 1). Six patients were previously hospitalized with COVID-associated respiratory distress, and one had received home-based oxygen therapy due to the unavailability of hospital beds at his home place. These seven patients also had documented history of steroid intake for an average of one week.

**Table 1:** Basic characteristics and clinical presentations of ten COVID-19 associated mucor-aspergillosis patients

| Case no. | Clinical presentation | Interval between symptoms related to COVID-19 and to fungal infection (in days) | Comorbidities/ immuno-compromised state | Other risk factors | HbA1C (%) | Procalcitonin (ng/mL) | Initial diagnosis |
| --- | --- | --- | --- | --- | --- | --- | --- |
| 1 | Headache, fever, altered sensorium, decrease vision | 15 | Uncontrolled Diabetes mellitus x 3 year | Severe COVID pneumonia; Smoker 10 pack years, H/O Steroid, low dose, > 1 week | 8.7 | 3.77 | Probable Rhino orbital mucormycosis with COVID 19<br>Probable Aspergillosis |
| 2 | Headache, Right nasal obstruction, rt. periorbital swelling, right eye tear with black nasal discharge | Not tested before admission (COVID symptoms-25 days back) | Uncontrolled Diabetes mellitus x 3 year | Severe COVID pneumonia. H/O Steroid intake for 5 -7 days; home- based Oxygen therapy Smoker H/O exposure to saw-dust | 11.7 | 0.07 | Proven Rhino orbital mucormycosis with COVID 19<br>Proven Aspergillosis |
| 3 | Headache, facial swelling, Fever | 22 | Newly diagnosed uncontrolled diabetes mellitus | Severe COVID pneumonia; H/O Non-invasive ventilation (Bi-PAP) for 20 days with high dose parenteral steroid > 2 weeks | 9.8 | NA | Proven RhinoorbitalMucormycosis<br>Probable Aspergillosis |
| 4 | Facial pain, nasal stuffiness, nasal discharge | 10 | Diabetes mellitus x 10 year | Moderate to severe COVID pneumonia. H/O Oxygen therapy and high steroid intake for 5-7 days; | 9.3 | 11.94 | Probable Sino nasal mucor mycosis<br>Probable Aspergillosis |
| 5 | Headache, swelling, nasal discharge, right eye pain | Not tested before admission (H/O fever with COVID-19 symptoms 20 days back) | Newly diagnosed uncontrolled diabetes mellitus | Mild COVID, Incision & drain in hard palate 10 days before presentation, unsterile dressing?? Open wound | 9.4 | 0.08 | Proven Rhino orbital mucormycosis with COVID 19<br>Proven Aspergillosis |
| 6 | Eye pain, swelling, facial numbness | Not tested before admission (H/O fever with cough, 10 days prior to admission) | Diabetes mellitus x 7 years | Moderate COVID, H/O low dose Steroid intake<br>Chronic kidney disease; H/O Dialysis;<br>Smoker | 5.8 | 0.89 | Proven Rhino orbital mucormycosis with COVID 19<br>Proven Aspergillosis |
| 7 | Headache, periorbital swelling, vomiting | Not tested before admission (Admitted to other centre with symptoms suggestive of COVID-19, H/O fever with cough, 11 days) | Uncontrolled Diabetes mellitus x 4 years, CKD | Mild COVID, Smoker | 13.4 | 0.05 | Proven Rhinoorbital Mucormycosis<br>Probable Aspergillosis |
| 8 | Headache, facial numbness, swelling, nasal discharge | 16 | Uncontrolled Diabetes mellitus x 4 years | Moderate COVID, Previous hospitalization for COVID-19 for 1 week, H/O High dose Steroid intake | 9.4 | 0.11 | Proven Rhino orbital mucormycosis with COVID 19<br>Proven Aspergillosis |
| 9 | Right eye swelling, pain, discharge | 5 | Diabetes mellitus x 1 years | Severe COVID pneumonia, No significant other history | 15.1 | NA | Proven Rhino orbital mucormycosis with COVID 19<br>Proven Aspergillosis |
| 10 | fever, cough, breathlessness, swelling of nose | 23 | Newly diagnosed diabetes mellitus; Pancytopenia | Severe COVID pneumonia, H/O High dose Steroid intake; H/O Blood transfusion (5 whole units); RECOVERED COVID PNEUMONITIS (2/5/21 – 19/5/21) | 6.3 | 3.78 | Probable Nasal Mucormycosis<br>Probable Aspergillosis |

**Figure 1:**
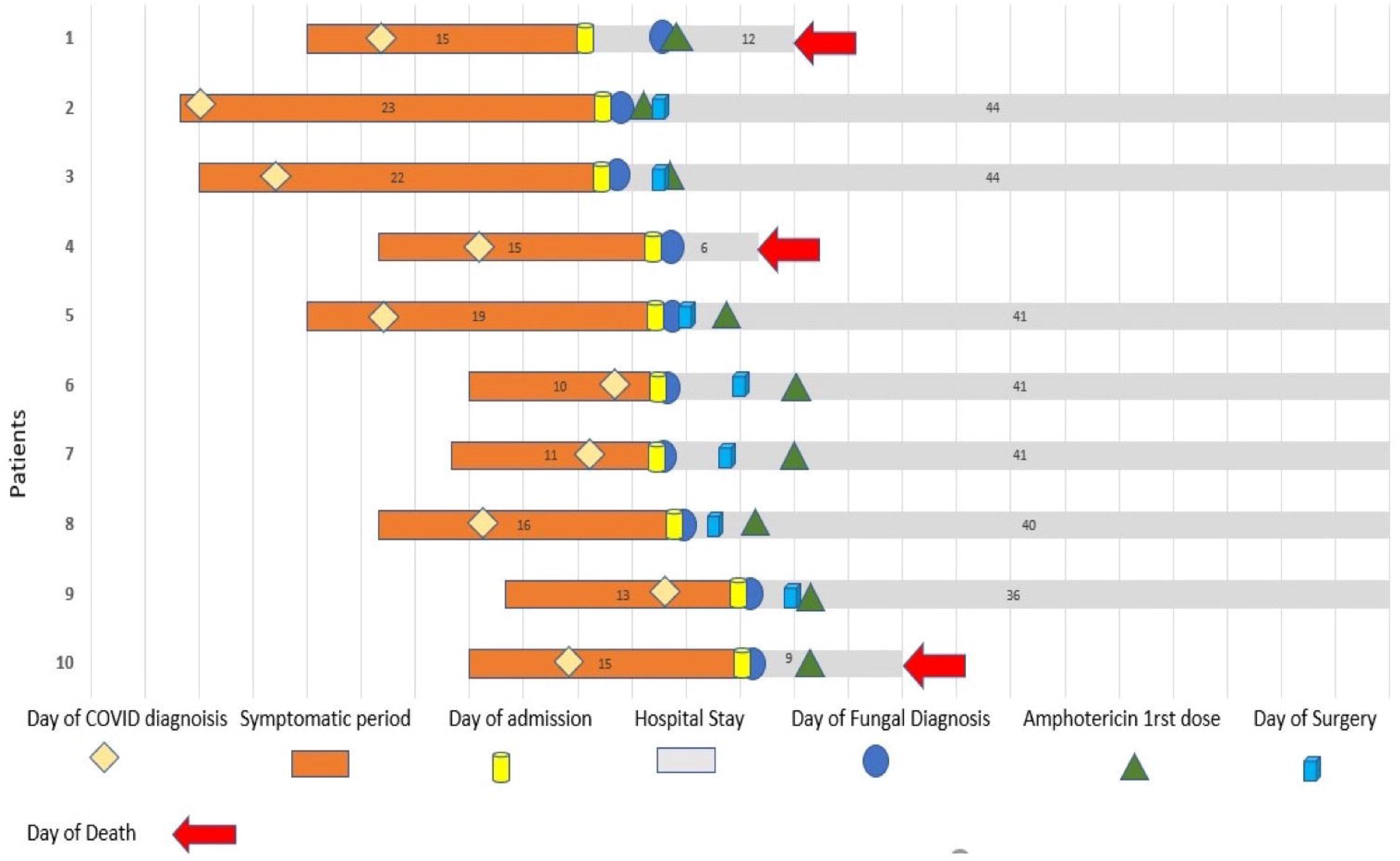
Schematic timeline presentation from day of symptom onset to outcome for 10 COVID-19 associated mucor-aspergillosis patients.

Patient 1 was diagnosed as a case of diabetic ketoacidosis with severe COVID-associated pneumonia and sepsis. On day-5 and day-9, two consecutive nasal tissue samples were sent to the laboratory for KOH mount that showed acute angle branched thin hyaline septate hyphae, suggestive of *Aspergillus* spp. and was started on injection amphotericin-B. The patient’s condition worsened, on day-14 repeat nasal tissue KOH revealed both broad aseptate perpendicular branching hyphae and thin hyaline septate hyphae. But the patient succumbed a day later.

Pre-operative nasal tissue KOH stain of Patient 2 was suggestive of *Mucorales* and *Aspergillus* whereas only broad aseptate hyphae were visible in the post-operative sample, but the fungal culture of both samples showed growth of *R. arrhizus* and *A. flavus*.

Patients 4 and 10 were intubated and managed conservatively by medical management only. Patients 3,7,8, and 9 were diagnosed on the same day of their admission and were managed with medical therapy aided by endoscopic debridement. On follow-up, these patients are recovering well to date.

Radiological, microbiological, and histopathological (HPE) diagnosis, treatment, and outcome of the patients were described (Table 2). Nasal tissue of all patients showed growth of *R. arrhizus* with *A. flavus* in seven and *A. fumigatus* in three patients. Other laboratory markers were described in supplementary table 1. Only 70% of patients (7/10) were managed successfully in terms of saving a life.

**Table 2:**
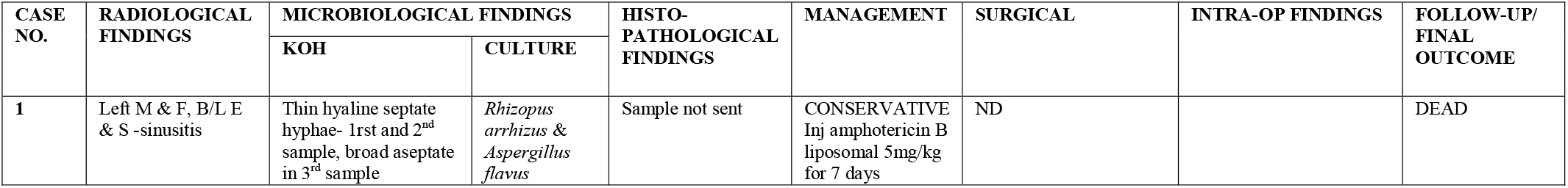
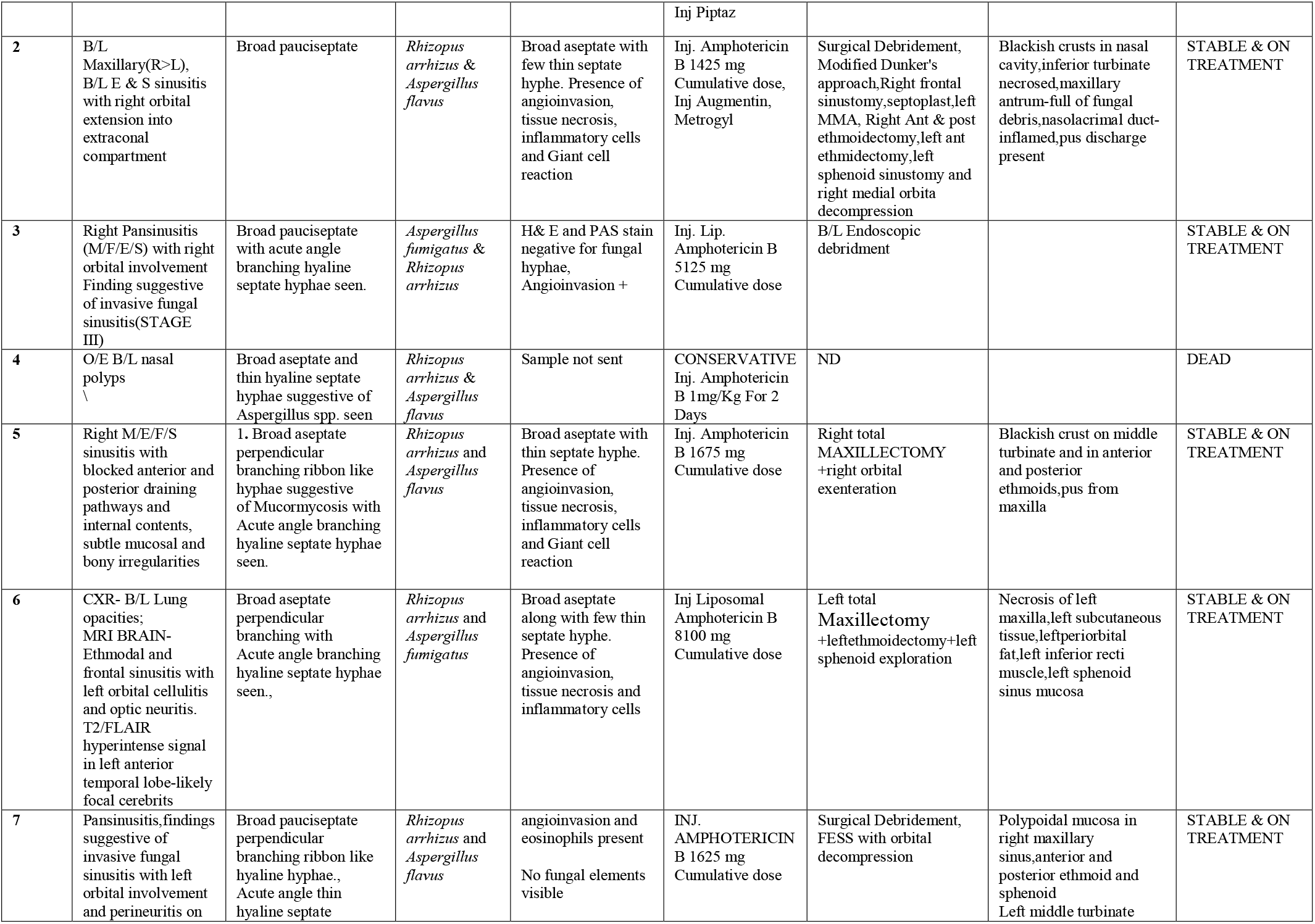
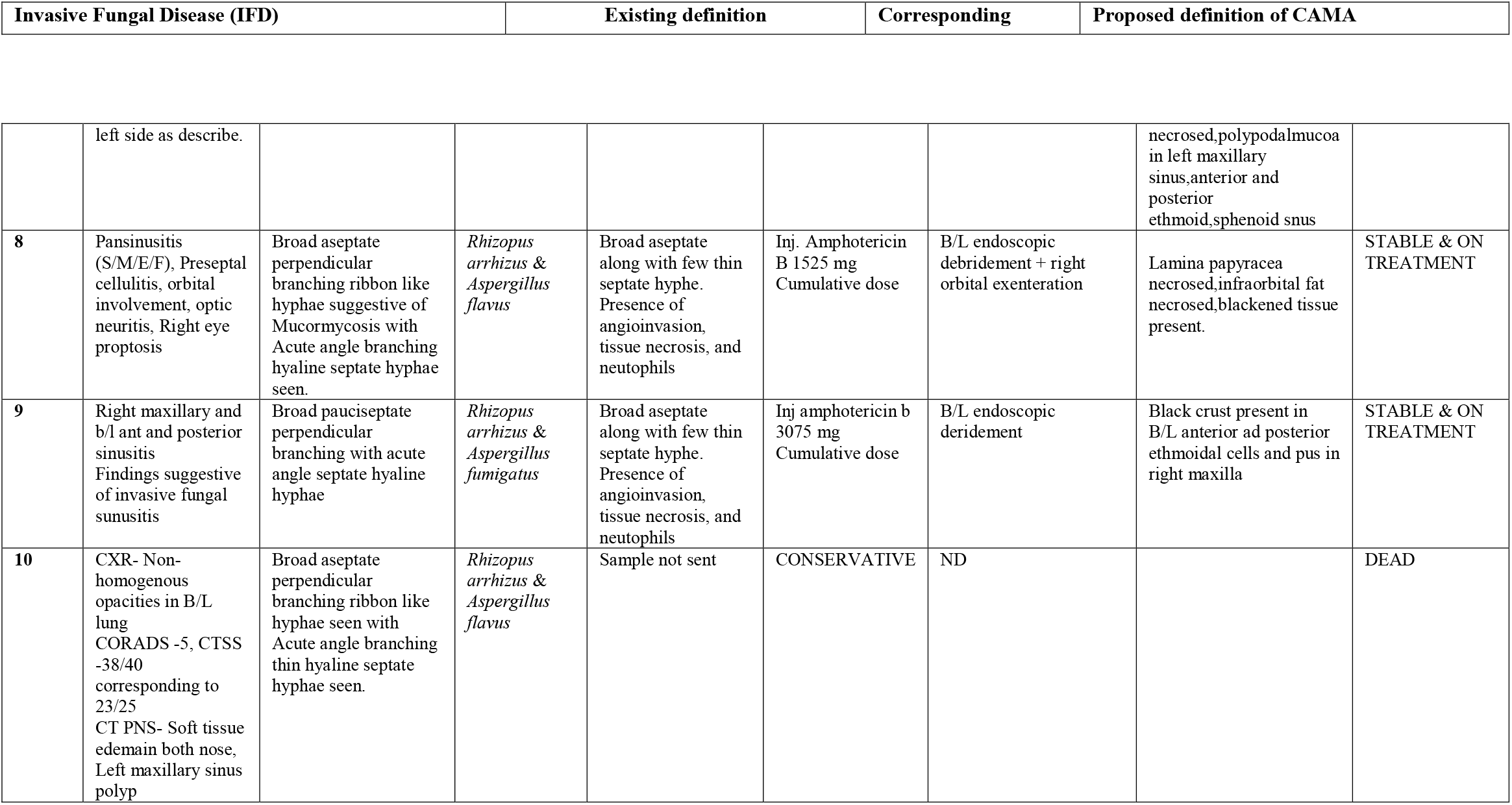
Radiological and microbiological diagnosis, treatment, and outcome of ten COVID-19 associated mucor-aspergillosis patients

## DISCUSSION

During the first wave of COVID, the primary focus was on COVID–associated pulmonary aspergillosis (CAPA), other fungal superinfections, including *Candida* infections and rare mold infections, whereas the second wave of COVID pandemic, especially in India, has observed an exponential increase of cases of COVID–associated mucormycosis (CAM) [6,11-13]. Rapid diagnosis of secondary infections and co-infections is complex and crucial for early management. Mucormycosis is a life-threatening entity however, the true impact of *Aspergillus* co-infections remains unknown. There is a paucity of data regarding mixed mucormycosis and aspergillosis co-infection in the available literature with no such data available in COVID patients i.e. CAMA.

*Mucorales* and *Aspergillus* are ubiquitous soil-dwelling organisms, and humans are exposed daily to fungal elements. They dwell in decaying organic matter, soil, animal excreta, dust, compost, foods, spices, unfiltered air, and ventilation system. *Aspergillus* spp. is one of the most commonly isolated fungus among healthy volunteers and forms part of their mycobiota [14].

The most important risk factors for mucormycosis and aspergillosis include uncontrolled diabetes mellitus, and use of corticosteroids leading to hyperglycemia, extensive use of broad-spectrum antibiotics, prolonged hospitalization, and presence of co-morbidities such as structural lung defects, with diabetes being most common risk factor for mucormycosis [15]. Apart from aforementioned risk factors, cytokine storm activated by the viral antigens, high airway pressure, and hypoxia-induced acute lung injury secondary to mechanical ventilation along woth drug toxicity increases risk of invasive aspergillosis. Moreover, in ideal situations, immunocompromised patients should be kept in isolation rooms with positive pressure but to protect healthcare workers from COVID, it was recommended to keep COVID positive patients in negative pressure rooms. Ichai *et al*. also demonstrated that patients admitted in negative pressure ICU rooms were at higher risk of aspergillosis and other secondary infections as well [16]. Patient’s co-morbidities further influence the likelihood of secondary fungal infections [17]. In this study, patients 1, 6, and 7 had a history of cigarette smoking which is strongly associated with adverse outcomes from COVID in studies [18].

In this study, all patients suffered from COVID-associated pneumonitis either at admission or within the past one month suggesting that residual pathology from an initial infection by respiratory viral infections can directly damage airway epithelium and facilitate invasion of tissues by molds like *Mucorales* and *Aspergillus* spp [19]. SARS CoV-2 leads to immune dysfunction or dysregulation, leading to a decreased T-cell population, mostly affecting the CD4+ CD8+ T-cell subset and induces lymphocytopenia, thus giving way to bacterial and fungal superinfection [9,20]. In the case of hematological malignancies, severe lymphopenia has earlier been established as a predicting risk factor for invasive mold disease as was also observed in patient 10 in this study who had pancytopenia [21].

Although the triad of COVID pneumonia, diabetes, and steroid use is common for patients worldwide, yet secondary fungal infections have been exponentially reported from India. Few possible reasons for this may be environmental factors, widespread use of steroids, monoclonal antibodies, broad-spectrum antibiotics, antiparasitics, antivirals as part of cocktail therapy used against COVID even in mild cases with no strict prescription checks in India and most drugs including glucocorticoids readily available over the counter. The pre-COVID baseline prevalence of mucormycosis (14 cases/100,000 people) in India is much higher than in the rest of the world (0.2/100,000) [4]. In a multicentric study done in Indian ICUs, *Aspergillus* species were the commonest (82.1%) mold isolated [22]. What’s more, India is the diabetes capital of the world, having the second-largest population with diabetes globally and SARS CoV-2 virus has been found to be more prevalent and severe in people with diabetes [23, 24]. Hyperglycaemia incites hypervirulence of specific pathogens and is also an independent risk factor for bacterial and fungal co-infections [25].

Irrational use of steroids either for a long duration or during the early phase of COVID illness or in higher than recommended doses has been cited as one of the most common underlying causes for fungal superinfection in COVID patients [4].

Environmental factors play a significant role in increasing CAM and CAPA in the Indian subcontinent. The atmosphere is more conducive for the fungus to thrive in South Asian developing countries [26]. There are a few unanswered and unresolved theories like incessant use of industrial-grade oxygen and use of herbal products may have caused this increase in fungal diseases. There could also be a possible role of contaminated water used in humidifiers, unsterile humidifier bottles, and non-medical grade industrial oxygen cylinders in an increase of fungal infections through the spread of fungal spores [27]. Though fungal spores do not survive in water yet chances of transmission of spores may be there from high contact hospital surfaces including reusable humidifier bottles. Considering abovementioned viewpoints, the present study patients also had such exposures, but a direct relationship couldn’t be established.

Patient 5 in this study had a history of incision and drainage for dental extraction at a local clinic suggesting her case likely to be healthcare-associated. Few patients in this study also had a history of previous hospitalization, which may have exposed patients to fungal infections. Studies have reported such secondary fungal infections due to adhesive bandages, wooden tongue depressors, negative pressure rooms, poor air filtration, non-sterile medical devices water leaks, and building construction [28].

Diagnosis of fungal infections is often challenging due to their ubiquitous nature. Direct microscopy (10% KOH mount) and fungal cultures, although have low sensitivity (<50%), remain essential tools. Positive culture growth may often reflect laboratory contamination or colonization of nasal tissue and upper respiratory samples, rather than the true disease. HPE, though, may confirm invasiveness but is time-taking. Further studies must target to identify biomarkers indicating tissue invasion stages and airway involvement. Off lately, new molecular platforms are being investigated. *Mucorales* PCR in combination with screening assays for *Aspergillus* like serum galactomannan (GM) and (1→3)-β-d-glucan (BG) antigen assays are useful and should be implemented in high-risk cases [29]. Clinical and radiological findings need to be correlated with culture findings [8]. Culture sensitivity has little value in dual infections except identifying non-amphotericin antifungal identification if any to reduce amphotericin-B resistance in the future since after this drug no other broad-spectrum exists at present.

Based on this study findings, herein we propose case definitions for CAMA (Table 3). Since, the included cases had mixed infection,they were presumed to be proven if both fungal type were of proven type. However, in reality patients may have proven mucor with probable or possible aspergillosis or any combinations. Then what to do?

**Table 3:** Proposed Case definitions for COVID-19 associated mucor-aspergillosis (CAMA) patients

| (EORCT/MSG Criteria) |  |  | Present case series as per existing combined definition |  |
| --- | --- | --- | --- | --- |
|  | MUCORMYCOSIS | ASPERGILLOSIS | CAMA |  |
| **PROVEN IFD:**<br>1. **Host factors:** Any patients<br>2. Confirmed with Histopathologic, cytopathologic, or direct microscopic examination of a specimen or culture of a specimen of tissue taken from a site of disease; obtained from a normally sterile and clinically or radiological abnormal site consistent with an infectious disease process | Corresponds | Corresponds | Cases 2,3,5,6,7,8,9 | Corresponds, but in combination of IFD cases such as CAMA one IFD may be proven and another probable type. |
| **PROBABLE IFD:**<br>presence of at least 1 host factor, a clinical feature and mycologic evidence, proposed for immunocompromised patients only.<br>1. **Host factors** (one of the following): Recent history of neutropenia (500 neutrophils/mm<sup>3</sup>) for 110 d, Receipt of an allogeneic stem cell transplant, Prolonged use of corticosteroids at a mean minimum dose of 0.3 mg/kg/d of prednisone equivalent for 13 wk, Treatment with other recognized T-cell immunosuppressants, Inherited severe immunodeficiency<br>2. **Clinical features** (one of the following three signs on CT) Dense, well-circumscribed lesion(s) with or without a halo sign, Air-crescent sign, Cavity, Wedge-shaped and segmental or lobar consolidation<br>3. **Mycological evidence:**<br>**Direct test** (cytology, direct microscopy, or culture): Mold in sputum, BAL, bronchial brush, or sinus aspirate samples | Corresponds | Corresponds | Cases 1,4,10 | Corresponds, but additional host factors for Aspergillosis to be included:<br>I. Concurrent or recently treated COVID-19 (≤6 weeks)<br>II. Uncontrolled DM<br>III. CKD<br>IV. Workers or farmers<br>V. Asthma<br>VI. Cystic Fibrosis<br>VII. Environmental factors (construction sites)<br>VIII. Colonizer<br>Additional host factors for Mucormycosis to be included:<br>I. Concurrent or recently treated COVID-19 (≤6 weeks)<br>II. Uncontrolled DM<br>III. CKD<br>IV. Iron overload<br>V. Trauma<br>VI. Antifungal prophylaxis<br>VII. Intravenous drug use |
| **POSSIBLE:**<br>Presence of host factors and clinical features but in the absence of or negative mycological findings.<br>For immunocompromised patients only | Corresponds | Corresponds | - | Corresponds |

This study has its own limitations with lack of serum GM test which could have helped identifying the cases as of proven invasive aspergillosis. In the study, despite an early diagnosis of fungal infection post-admission, most participants could not be started on antifungal immediately with an average gap of 4-5 days in diagnosis and start of antifungal due to limited supply of amphotericin-B (Figure 1). Similar observations were made in a multicentric study conducted in other parts of India [30]. A delay of a week in diagnosis may double the 30-day mortality from 35% to 66% [3]. Treating CAMA is more worrisome since one (aspergillosis is a metastatic invasive infection) focuses majorly on medical treatment while the other focuses equally on surgical and medical management (mucor is a locally invasive infection). Patient 1 was diagnosed about 10 days after admission and couldn’t be saved, reflecting on the need for timely diagnosis and treatment.

In conclusion, mixed fungal infections i.e. CAMA as COVID sequelae may be an emerging disease. This outbreak was seen particularly in COVID (or post-) patients with uncontrolled diabetes, on steroids, or cocktail therapy, or living in unhygienic environments. So judicious and relevant use of various medicines (drug stewardship) along with strict maintenance of personal and environmental hygiene is the key to prevent CAMA in COVID era. Patients should be advised to keep a check on their glycaemic index and seek medical advice at the earliest. Despite the increased risk of fatality due to CAMA, active multidisciplinary teamwork may help reducing mortality by diagnosing and managing timely. The focus should be on empirical management: hit hard approach with both medical and surgical treatment. The study’s findings suggest that timely and accurate diagnosis of CAMA is a must for effective disease management and thus improve outcomes. CAMA may often remain unreported. Identification of risk factors for CAMA and measures to prevent the same may help manage invasive fungal infections in COVID patients, thus decreasing overall morbidity. Lastly, the criteria to classify into proven, possible, and probable types of IFD among CAMA patients need a change.

## Data Availability

It will be made available to others as required upon requesting the corresponding author.

## Contributors

VS, AP, and PKP: Conceptualization, Methodology, Data curation, Original draft preparation. MT, AT, and AT: Visualization, Investigation. PKP, AKS and SR: Supervision, critically reviewing. AT and MB: Reviewing and Editing, Validation. All authors approved the drafting.

## Data sharing

It will be made available to others as required upon requesting the corresponding author.

## Acknowledgment

We would like to thanks the COVID and MUCOR management team, AIIMS Rishikesh in managing all patients and contributing patient data documentation.

## Declarations

No funding was received to assist with the preparation of this manuscript. The authors have no conflicts of interest to declare that are relevant to the content of this article. This case series is part of a project titled disease profile of COVID-l9 (DPC-I9), including their follow-up in patients at a tertiary institute in India, approved by the Institutional Ethical Committee, All India Institute of Medical Sciences (AIIMS), Rishikesh.

## REFERENCES

1. Hu B, Guo H, Zhou P, Shi ZL. Characteristics of SARS-CoV-2 and COVID-19. Nat Rev Microbiol. 2021;19(3):141–154. https://doi.org/10.1038/s41579-020-00459-7

2. Cevik M, Kuppalli K, Kindrachuk J, Peiris M. Virology, transmission, and pathogenesis of SARS-CoV-2. BMJ. 2020;371:m3862. https://doi.org/10.1136/bmj.m3862

3. Werthman-Ehrenreich A. Mucormycosis with orbital compartment syndrome in a patient with COVID-19. Am J Emerg Med. 2021 Apr;42:264.e5–264.e8; https://doi.org/10.1016/j.ajem.2020.09.032.

4. Moorthy A, Gaikwad R, Krishna S, Hegde R, Tripathi KK, Kale PG, Rao PS, Haldipur D, Bonanthaya K. SARS-CoV-2, Uncontrolled Diabetes and Corticosteroids—An Unholy Trinity in Invasive Fungal Infections of the Maxillofacial Region? A Retrospective, Multi-centric Analysis. J Maxillofac Oral Surg. 2021 Mar 6:1–8; https://doi.org/10.1007/s12663-021-01532-1

5. Martin Hoenigl, Invasive Fungal Disease Complicating Coronavirus Disease 2019: When It Rains, It Spores, Clinical Infectious Diseases, 2020; ciaa1342; https://doi.org/10.1093/cid/ciaa1342

6. Prakash H, Chakrabarti A. Epidemiology of Mucormycosis in India. Microorganisms. 2021; 9(3):523; https://doi.org/10.3390/microorganisms9030523

7. Gangneux JP, Bougnoux ME, Dannaoui E, Cornet M, Zahar JR. Invasive fungal diseases during COVID-19: We should be prepared. J Mycol Med. 2020;30(2):100971; https://doi.org/10.1016/j.mycmed.2020.100971

8. Cornely OA, Alastruey-Izquierdo A, Arenz D, Chen SC, Dannaoui E, Hochhegger B, Hoenigl M, Jensen HE, Lagrou K, Lewis RE, Mellinghoff SC. Global guideline for the diagnosis and management of mucormycosis: an initiative of the European Confederation of Medical Mycology in cooperation with the Mycoses Study Group Education and Research Consortium. The Lancet infectious diseases. 2019 Dec 1;19(12):e405–21; https://doi.org/10.1016/S1473-3099(19)30312-3. Epub 2019 Nov 5.

9. Koehler P, Bassetti M, Chakrabarti A, et al. Defining and managing COVID-19-associated pulmonary aspergillosis: the 2020 ECMM/ISHAM consensus criteria for research and clinical guidance. Lancet Infect Dis. 2021;21(6):e149–e162; https://doi.org/10.1016/S1473-3099(20)30847-1

10. □□□□ □□□□□□ □□ □ □□□□ □□□□□ □□ □□□□□, □□□□ □□□□□□ □ [Internet]. [cited 2021 Jun 12]. Available from: https://aiimsrishikesh.edu.in/a1_1/wp-content/uploads/2020/11/MUCOR-Management-protocol_AIIMS-Rishikesh_Version-1.0.cleaned.pdf

11. Arastehfar A, Carvalho A, van de Veerdonk FL, Jenks JD, Koehler P, Krause R, Cornely OA, S Perlin D, Lass-Flörl C, Hoenigl M. COVID-19 associated pulmonary aspergillosis (CAPA)—from immunology to treatment. Journal of Fungi. 2020 Jun;6(2):91; https://doi.org/10.3390/jof6020091.

12. Bartoletti M, Pascale R, Cricca M, et al. Epidemiology of invasive pulmonary aspergillosis among COVID-19 intubated patients: a prospective study [published online ahead of print, 2020 Jul 28]. Clin Infect Dis. 2020;ciaa1065; https://doi.org/10.1093/cid/ciaa1065

13. John, T.M.; Jacob, C.N.; Kontoyiannis, D.P. When Uncontrolled Diabetes Mellitus and Severe COVID-19 Converge: The Perfect Storm for Mucormycosis. J. Fungi 2021, 7, 298; https://doi.org/10.3390/jof7040298

14. Krüger W, Vielreicher S, Kapitan M, Jacobsen ID, Niemiec MJ. Fungal-Bacterial Interactions in Health and Disease. Pathogens (Basel, Switzerland). 2019 May;8(2); https://doi.org/10.3390/pathogens8020070.

15. Cox MJ, Loman N, Bogaert D, O’Grady J. Co-infections: potentially lethal and unexplored in COVID-19. Lancet Microbe. 2020;1(1):e11; https://doi.org/10.1016/S2666-5247(20)30009-4.

16. Ichai P, Saliba F, Baune P, Daoud A, Coilly A, Samuel D. Impact of negative air pressure in ICU rooms on the risk of pulmonary aspergillosis in COVID-19 patients. Crit Care. 2020;24(1):538. Published 2020 Sep 1; https://doi.org/10.1186/s13054-020-03221-w

17. Cimolai N. The Complexity of Co-Infections in the Era of COVID-19 [published online ahead of print, 2021 Apr 23]. SN Compr Clin Med. 2021;1–13; https://doi.org/10.1007/s42399-021-00913-4.

18. Patanavanich R, Glantz SA. Smoking Is Associated With COVID-19 Progression: A Meta-analysis. Nicotine Tob Res. 2020;22(9):1653–1656; https://doi.org/10.1093/ntr/ntaa082

19. Pittet LA, Hall-Stoodley L, Rutkowski MR, Harmsen AG. Influenza virus infection decreases tracheal mucociliary velocity and clearance of Streptococcus pneumoniae. Am J Respir Cell Mol Biol. 2010;42(4):450–460; https://doi.org/10.1165/rcmb.2007-0417OC

20. Kothari A, Singh V, Nath UK, et al. Immune Dysfunction and Multiple Treatment Modalities for the SARS-CoV-2 Pandemic: Races of Uncontrolled Running Sweat?. Biol (Basel). 2020;9(9):243; https://doi.org/10.3390/biology9090243

21. Magira EE, Jiang Y, Economides M, Tarrand J, Kontoyiannis DP. Mixed mold pulmonary infections in haematological cancer patients in a tertiary care cancer centre. Mycoses. 2018;61(11):861–867; https://doi.org/10.1111/myc.12830

22. Chakrabarti A, Kaur H, Savio J, et al. Epidemiology and clinical outcomes of invasive mould infections in Indian intensive care units (FISF study). J Crit Care. 2019;51:64–70; https://doi.org/10.1016/j.jcrc.2019.02.005

23. Mohan V. Why are Indians more prone to diabetes?. J Assoc Physicians India. 2004;52:468–474.

24. Huda MSB, Shaho S, Trivedi B, et al. Diabetic emergencies during the COVID-19 pandemic: A casecontrol study. Diabet Med. 2021;38(1):e14416; https://doi.org/10.1111/dme.14416.

25. Wang S, Ma P, Zhang S, et al. Fasting blood glucose at admission is an independent predictor for 28-day mortality in patients with COVID-19 without previous diagnosis of diabetes: a multi-centre retrospective study. Diabetologia. 2020;63(10):2102–2111; https://doi.org/10.1007/s00125-020-05209-1.

26. Thompson Iii GR, Cornely OA, Pappas PG, et al. Invasive Aspergillosis as an Under-recognized Superinfection in COVID-19. Open Forum Infect Dis. 2020;7(7):ofaa242; https://doi.org/10.1093/ofid/ofaa242

27. Banerjee M, Pal R, Bhadada SK. Intercepting the deadly trinity of mucormycosis, diabetes and COVID-19 in India [published online ahead of print, 2021 Jun 8]. Postgrad Med J. 2021;postgradmedj-2021-140537; https://doi.org/10.1136/postgradmedj-2021-140537

28. Rammaert B, Lanternier F, Zahar JR, et al. Healthcare-associated mucormycosis. Clin Infect Dis. 2012;54 Suppl 1:S44–S54; https://doi.org/10.1093/cid/cir867

29. Guegan H, Iriart X, Bougnoux ME, Berry A, Robert-Gangneux F, Gangneux JP. Evaluation of MucorGenius® mucorales PCR assay for the diagnosis of pulmonary mucormycosis. J Infect. 2020;81(2):311–317; https://doi.org/10.1016/j.jinf.2020.05.051

30. Patel A, Agarwal R, Rudramurthy SM, et al. Multicenter Epidemiologic Study of Coronavirus Disease-Associated Mucormycosis, India [published online ahead of print, 2021 Jun 4]. Emerg Infect Dis. 2021;27(9):10.3201/eid2709.210934; https://doi.org/10.3201/eid2709.210934

